# Immune response induced by the Recombinant Novel Coronavirus Vaccine (Adenovirus Type 5 Vector) (Ad5-nCoV) in persons living with HIV (PLWH)

**DOI:** 10.1101/2024.10.17.24315660

**Authors:** P. Cahn, L. Barreto, M.I. Figueroa, V. Fink, M.J. Rolón, G. Lopardo, I. Casetti, M. Ceschel, P. Patterson, L. Gambardella, G. Miernes, A. Nava, J. Gou, R. Wang, T. Zhu, S.A. Halperin

## Abstract

**Background:** Despite higher risk of poorer outcomes and potential sub-optimal vaccine effectiveness, people living with HIV (PLWH) are underrepresented in SARS-CoV-2 vaccine trials. We evaluated the safety and immunogenicity of the Ad5-nCoV vaccine (CanSino Biologics Inc./The Beijing Institute of Biotechnology) in PLWH.

**Methods:** In this single arm, open-label Phase 2b trial, PLWH were enrolled in Argentina. Participants received two doses of Ad5-nCoV vaccine (intramuscular, dosage 5×10^10^ viral particles) at days 0 and 56. The primary outcomes were safety as serious adverse events [SAE], solicited and unsolicited local and systemic adverse events, impact on HIV viral load and CD4 counts and immunogenicity measured by S-RBD IgG and pseudo-virus neutralizing antibodies (nAbs) up to 52 weeks (ClinicalTrials.gov:NCT05005156).

**Findings:** Between June 2021-January 2022, 140 PLWH received at least one dose of Ad5-nCoV vaccine. At baseline, the majority were on antiretroviral therapy (99.3%), virologically suppressed (93.6%), with a median (IQR) CD4-cell count:736 (531-946) cells/ul. At baseline, 38 (27%) participants were seropositive for S-RBD antibodies, and 40 (28%) for nAbs. There were no SAEs related to the vaccine. Solicited AE within 7 days after first and second dose occurred in 93 (69%) and 75 (60%) participants, mostly grade 1, included pain, drowsiness and headache. The incidence of unsolicited AE within 28 days of vaccination was 10.7%. There were no significant changes in plasma viral load, CD4, CD4/CD8 ratio and no new AIDS-defining illnesses were reported. There were significant increases in the geometric mean titers (GMT) of S-RBD and nAbs between baseline to week 52. Seroconversion rates 28 days after the first and second doses (day 84) were 80% and 94% for S-RBD, and 35% and 78% for nAbs.

**Interpretation:** Two doses of the Ad5-nCoV vaccine were safe and induced an adequate immune response in virologically suppressed PLWH, maintaining high antibody titers at least during the first year post-vaccination.

**Funding:** CanSino Biologics Inc, Tianjin, China

## Introduction

In March 2020, the World Health Organization declared a global pandemic caused by SARS-CoV-2 (COVID-19). Four years later (March 2024), there have been more than 774 million reported cases and more than 7 million deaths [1]. People living with HIV (PLWH) are at increased risk of worse clinical outcomes after COVID-19 infection, particularly those with lower CD4-cell counts or not receiving antiretroviral therapy (ART)[2,3].

Vaccines have played a key role in the public health response to the COVID-19 pandemic. Indeed, shortly after the declaration of the global COVID-19 pandemic, a number of vaccines were rapidly developed and proved to be safe and efficacious in preventing symptomatic COVID-19[4]. In addition, despite lower vaccine effectiveness against variants of concern, their protection against severe COVID-19 has remained high[5].

Despite higher risk of poorer outcomes and potential sub-optimal vaccine effectiveness, PLWH have been largely underrepresented in pivotal COVID-19 vaccine trials. PLWHIV represented only 1% (1,557) out of a total number of approximately 150,000 participants enrolled in phase 2/3 trials evaluating the mRNA-1273/Moderna, BNT162b2/Pfizer-BioNTech, ChAdOx1/AstraZeneca, NVX-2373/Novavax and Ad26.COV2.S/Janssen vaccines, with other studies completely excluding them[6].

To our knowledge, only three trials aimed to address concerns of a potential reduced immune response to COVID-19 vaccines among PLWH: two assessed the ChAdOx1/AstraZeneca vaccine[7,8] and one the NVX-2373/Novavax vaccine[9]. In these studies, two doses of COVID-19 vaccines were found to be safe and immunogenic among PLWH, particularly among SARS-CoV-2 baseline-seropositive participants. Despite these promising results, sample sizes were small (ranging from 52 to 122), only evaluated two types of vaccines and were conducted in only two countries (UK and South Africa), highlighting the need for additional high-quality experimental studies in PLWH. This is particularly important for SARS-CoV-2 vaccines based on adenovirus type-5 (Ad5) platforms, given prior concerns associated with higher risk of HIV acquisition in participants of Ad5 vectored HIV-1 vaccine trials[10]. Although follow-up data from one of these studies did not find increased risk of HIV progression among participants who became HIV-infected[11], it remains critical to monitor the effects of other Ad-5-based vaccines on HIV infection and progression. Therefore, the objective of this study was to assess the safety and immunogenicity of two doses of Ad5-nCoV/ CanSino Biologics Inc./The Beijing Institute of Biotechnology in PLWH.

## Methods

### Study design

The original protocol (version 1.0) included a 52-week randomized placebo-controlled clinical trial evaluating the safety and immunogenicity of one dose of the Ad5-nCoV vaccine in PLWHIV on stable ART and virologically suppressed. The rapid advancement of the Argentinean COVID vaccination program reduced the possibilities of recruiting the study under such design. Accordingly, the study was amended on October 6^th^, 2021 to change the study design to a Phase IIb, multicentre, open-label, single-arm trial evaluating the safety and immunogenicity of two doses of the Ad5-nCoV vaccine in PLWHIV (version 1.5). Exclusion criteria based on plasma HIV viral load or ART status were also removed. The trial was conducted in four sites in Buenos Aires, Argentina between May 31, 2021, and January 13, 2023, in accordance with the Declaration of Helsinki and Good Clinical Practice guidelines. All participants gave their written informed consent. The study was approved by ANMAT (Argentina’s regulatory agency) and by the Institutional Review Boards at each participating site, and is registered in clinicaltrials.gov: NCT05005156.

### Participants

We enrolled clinically stable adults (18 years and older) living with HIV who were free from opportunistic infections or AIDS-defining illness for at least six months before screening. Participants were excluded if they had a history of laboratory-confirmed SARS-CoV-2 infection, receipt of an adenovirus-vectored, coronavirus, or SARS-CoV-2 vaccine, any AIDS-defining illness, any medical or psychiatric unstable condition (including immunosuppression conditions other than HIV) that in the opinion of the investigator could preclude safe participation in the study, current substance use disorder or history of anaphylaxis to any vaccine component. Women who were pregnant or breastfeeding were not eligible to participate, and all participants involved in heterosexual sexual activity had to agree to the use of approved contraceptive methods.

### Procedures

Eligible and consenting participants received two standard 0.5 mL/vial (i.e., 5×10^10^vp) intramuscular doses of the Ad5-nCoV vaccine in the deltoid region given eight weeks apart (i.e., day 0 and 56), administered by trained study personnel. The Ad5-nCoV vaccine was produced as previously described[12].

All participants were contacted on a weekly basis (email, telephone, text message) to assess for any symptoms of COVID-19. The presence of any possible COVID-19 symptoms triggered laboratory testing for SARS-CoV-2 infection via PCR.

Participants were also asked to complete electronic or paper diaries reporting solicited adverse events (AEs) for 7 days and unsolicited AEs for 28 days after both the first and second vaccination. Serious adverse events (SAE) and medically attended adverse events (MAAE) were collected throughout the 52-week duration of the trial and recorded every 4 weeks via telephone calls.

A baseline SARS-CoV-19 antibody, and a urine pregnancy test in women of child-bearing potential was performed on Day 0, immediately before vaccination. Blood samples for immunogenicity analysis were collected on days 0, 28, 84, and weeks 24 and 52. Immunogenicity measures included IgG antibodies to the receptor-binding domain of the SARS-CoV-2 spike protein 9 (S-RBD IgG antibody), measured by ELISA-based assays and neutralising antibodies against SARS-CoV-2 (nAbs), measured by pseudovirus neutralisation assays. HIV viral load, CD4 and CD8-cell counts were measured on day 0, and weeks 24 and 52.

### Outcomes

The two co-primary outcomes were the safety and immunogenicity profile of the Ad5-nCoV vaccine. Safety was assessed by the incidence of solicited and unsolicited AE occurring within 7 and 28 days of each vaccination, respectively, SAE and MAAE throughout the study period, as well as HIV viral load at weeks 24 and 52. Immunogenicity was evaluated by the seroconversion rate of the S-RBD IgG antibody on Day 28, Day 84, Week 24, and Week 52 after vaccination.

Secondary safety outcomes included the incidence of a decrease in CD4-cell count by >=20% as well as the change of the CD4/CD8 ratio from the baseline value. Secondary immunogenicity outcomes included the seroconversion rate of nAbs, as well as the geometric mean titer (GMT) and geometric mean increase (GMI) of the S-RBD IgG antibody and nAbs at Day 28, Day 84, and Week 24 and Week 52 after vaccination.

Exploratory outcomes included the efficacy of two doses of Ad5-nCoV in preventing PCR-confirmed symptomatic COVID-19 occurring 14 and 28 days after the second dose (i.e., days 70 and 84) up to last study visit.

### Statistical analysis

Given that this was a non-comparative study; no formal sample size calculation was performed. The safety analysis population included all participants who received at least one dose of the Ad5-nCoV vaccine. Separate safety analyses were conducted for the first and second doses. Safety endpoints were summarized using frequency counts and percentages. The immunogenicity analysis population included participants with at least one dose of the vaccine and available antibody results. GMT, GMI, and seroconversion rate (with corresponding 95% confidence intervals) were calculated for each time point, using mixed effects linear regression models to assess changes over time. The seroconversion rate was calculated as the proportion of participants that had a GMI of the S-RBD IgG antibody or nAbs equal to or greater than 4 from baseline at each pre-specified time point. At baseline, participants with S-RBD IgG antibody or nAbs titers greater than 4 were considered positive. The evaluation of changes in the proportion of participants with undetectable VL, log10 VL, CD4/CD8-cell counts and CD4/CD8 ratio was performed fitting a mixed effects linear regression model, setting as fixed independent variable the time as a continuous variable, and subject as random term.

The analysis of the exploratory efficacy endpoint was descriptive and performed in the total participants that were vaccinated with two doses.

All statistical analysis was performed using R Statistical Software[13].

## Results

### Baseline characteristics

Between May 31, 2021 and January 6, 2022, 141 PLWHIV were enrolled in this study. Of these, 140 participants received at least one dose of the Ad5-nCoV vaccine (one participant withdrew consent before receiving any dose) and were included in this analysis: 133 participants received two doses and 7 participants one dose. Figure 1 shows the CONSORT flow diagram of participants and Table 1 summarizes the participants’ baseline sociodemographic and clinical characteristics. The median age was 41 (Interquartile range [IQR]: 31-48), 79.0% were males and 97.9% self-reported white race. All except one participant (n=139, 99.3%) were on ART, the majority (93.6%) were virologically suppressed (VL <40 copies/mL) and the median (IQR) CD4-cell count was 736 (531-946) cells/ul. At baseline, 38 (27%) participants were seropositive for S-RBD antibodies, and 39 (28%) were seropositive for nAbs.

**Figure 1.**
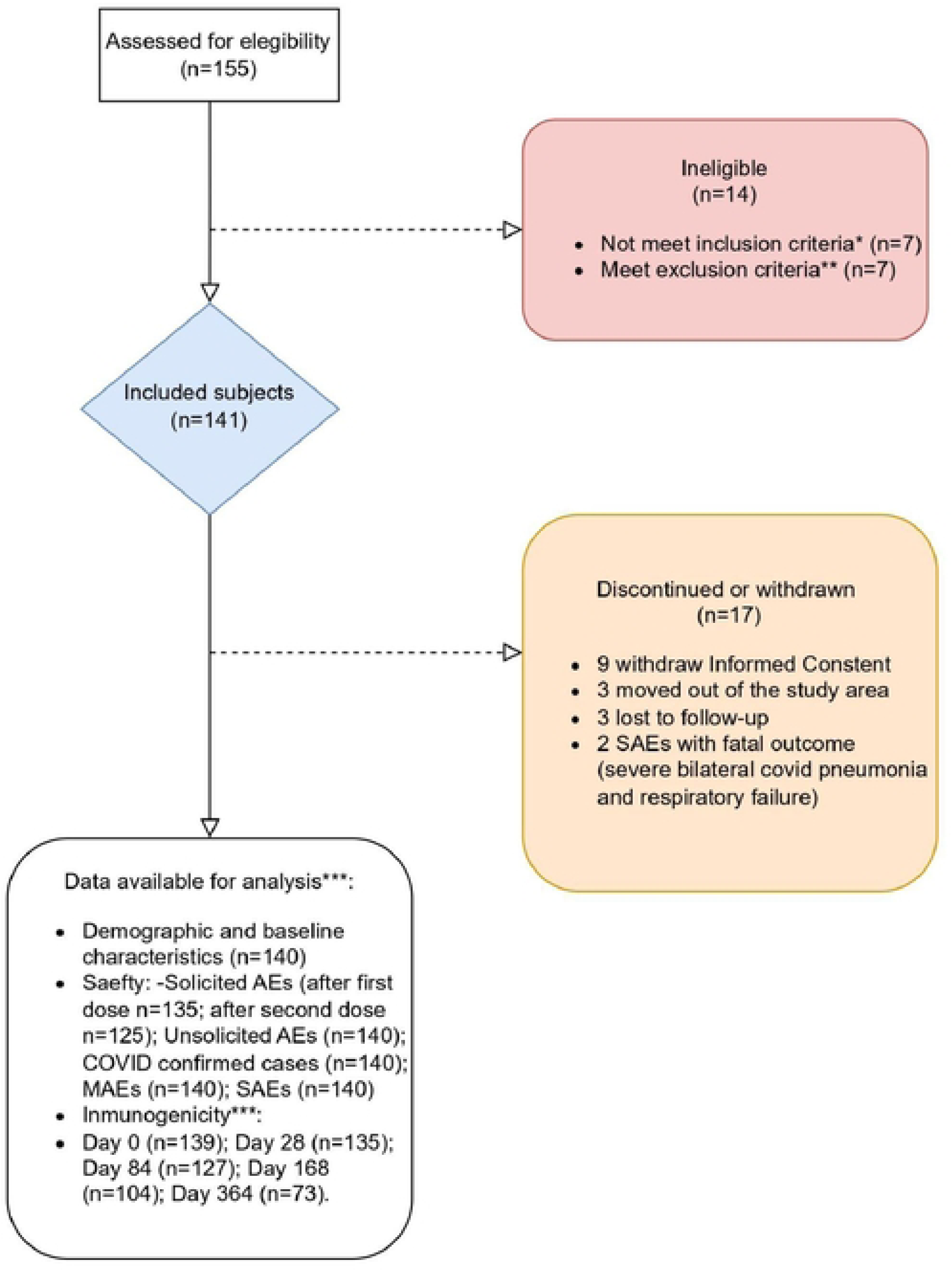
Consort Study Flow Chart.

**Table 1.** Baseline characteristics of the study population (N=140)

|  |  |
| --- | --- |
| Age - median (IQR*) | 41 (31-48) |
| Sex - % (95%CI) [n] |  |
| **Male** | **79% (70.7-84.9) [110]** |
| **Female** | **21% (15.1-29.3) [30]** |
| **Race - % (95%CI) [n]** |  |
| **White** | **97.9% (93.4-99.4) [137]** |
| **American Indian or Alaska Native** | **0.7% (0.4-4.5) [1]** |
| **Black** | **0.7% (0.4-4.5) [1]** |
| **Other** | **0.7% (0.4-4.5) [1]** |
| **on Antiretroviral therapy - % (95%CI) [n]** | **99.3% (97.9-100) [139]** |
| **Virologically suppressed /≤ 40 copies per mL- % (95%CI) [n]** | **93.6% (87.8-96.8) [131]** |
| **CD4 count (cells/ul) - median (IQR\*)** | **735.5 (530.5-945.8)** |

\*median (percentiles 25-75).

### Safety analysis

Solicited AE diaries were completed by 135 participants (96%) after the first dose, and 125 participants (94%) after the second dose. The proportion of participants reporting solicited systemic and local AE after the first and second dose are presented in Figure 2. After the first dose, 93 participants (69%) reported at least one solicited AE, mostly grade 1. Pain (48%) was the most common local solicited AE and drowsiness (39%) and headache (34%) the most common systemic solicited AE. After the second dose, 75 participants (60%) reported at least one solicited AE. A similar pattern of solicited AE was observed after the first and second dose though the proportion of participants reporting systemic solicited AE was lower (25% drowsiness, 23% headache).

**Figure 2.**
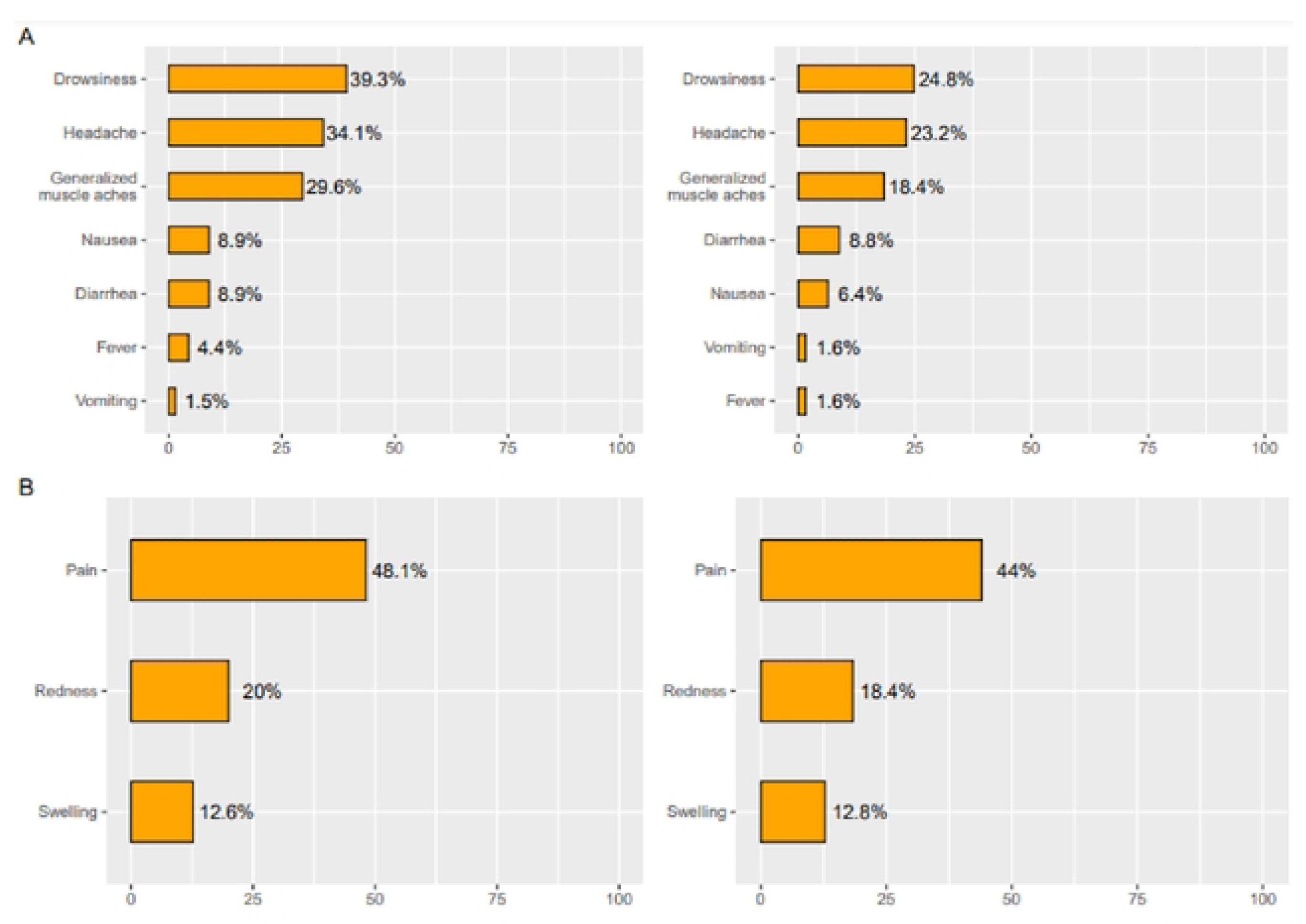
Proportion of participants reporting solicited systemic (A) and local (B) adverse events in participants vaccinated with at least one dose of the AdS-CoV vaccine, after first (left) and second (right) dose. The figure summarizes AEs at any day & grade up to 7 days after vaccination. Solicited AE diaries were completed by 135 participants after the first dose and 125 participants after the second dose.

The global incidence of unsolicited AE within 28 days of vaccination was 10.7% (15 events: 11 events after the first dose and 4 events after the second dose). The most frequent one was asthenia (7 events), only one was grade 3, and none required hospitalization.

Over the 52-week duration of the study, there were 8 SAEs (none of them related to the study vaccine). Two participants died, one due to myocardial infarction complicated with severe bilateral COVID pneumonia and the other due to fatal respiratory failure secondary to lung cancer.

A total of 141 MAAEs in 70 participants (50%) were reported, including 18 COVID-19 confirmed cases and 33 suspected cases. Most of the MAAEs were grade 2 (74%). Two MAAEs were possibly related to the study drug: acute unilateral hearing loss and fever of unknown origin.

HIV VL and CD4/CD8 cell count measurements at baseline, week 24 and 52 after the first vaccine dose are presented in Table 2. There were no significant changes in the proportion of participants who were virologically suppressed or the log10 VL in non-suppressed participants between baseline and the other time points. The mixed effects linear regression model fit showed a statistically significant increase of CD4, CD8-cell count mean, as well as CD4/CD8 ratio mean over time (p <0.001)[14].

**Table 2.** HIV viral load, CD4 and CD8 cell count measurements at baseline, week 24 and week 52 after the first Ad5-CoV vaccine dose.

|  | **Baseline**<br>**(n=140)** | **Week 24**<br>**(n=124)** | **Week 52**<br>**(n=123)** | **p-value** |
| --- | --- | --- | --- | --- |
| **Virologically**<br>**suppressed (<=**<br>**40 copies/ mL),**<br>**n (%)** | 131 (94) | 113 (91) | 112 (91) | 0.199 |
| **log10 HIV-VL**<br>**in non-**<br>**suppressed**<br>**participants,**<br>**median (IQR)** | 3.38 (2.37-4.59) | 2.38 (1.74-3.78) | 1.97 (1.77-3.46) | 0.062 |
| **CD4-cell count,**<br>**median (IQR)** | 735<br>(530.5-945.8) | 703.5<br>(515.8-940.5) | 789<br>(620-996.5) | **<0.001** |
| **CD8-cell count,**<br>**median (IQR)** | 886 (699-1144) | 833 (635.25-1059.5) | 1045<br>(751-1328) | **<0.001** |
| **CD4/CD8 ratio** | 0.80<br>(0.57-1.16) | 0.85<br>(0.60-1.14) | 0.82<br>(0.55-1.10) | **<0.001** |

A decrease of CD4-cell count of 20% or higher from baseline was observed in 22.6% of participants at week 24, and 12.2% at week 52. None of these participants had less than 200 CD4 cells per mL at any point in time, and there were no AIDS-defining illnesses during the observation period.

### Immunogenicity analysis

Immunogenicity analysis included samples from the 140 participants who received at least one vaccine dose and had available S-RBD antibodies and n-Abs results, excluding samples taken after COVID-19 confirmed cases: 139 samples for day 0, 135 for day 28, 127 for day 84, 104 for week 24, and 73 for week 52. Sixty-four samples were excluded due to SARS-CoV-2 confirmed infection. Tables 3 and 4 present GMT, GMI, and seroconversion rates at pre-specified time points for S-RBD antibodies and nAbs, respectively.

**Table 3.**
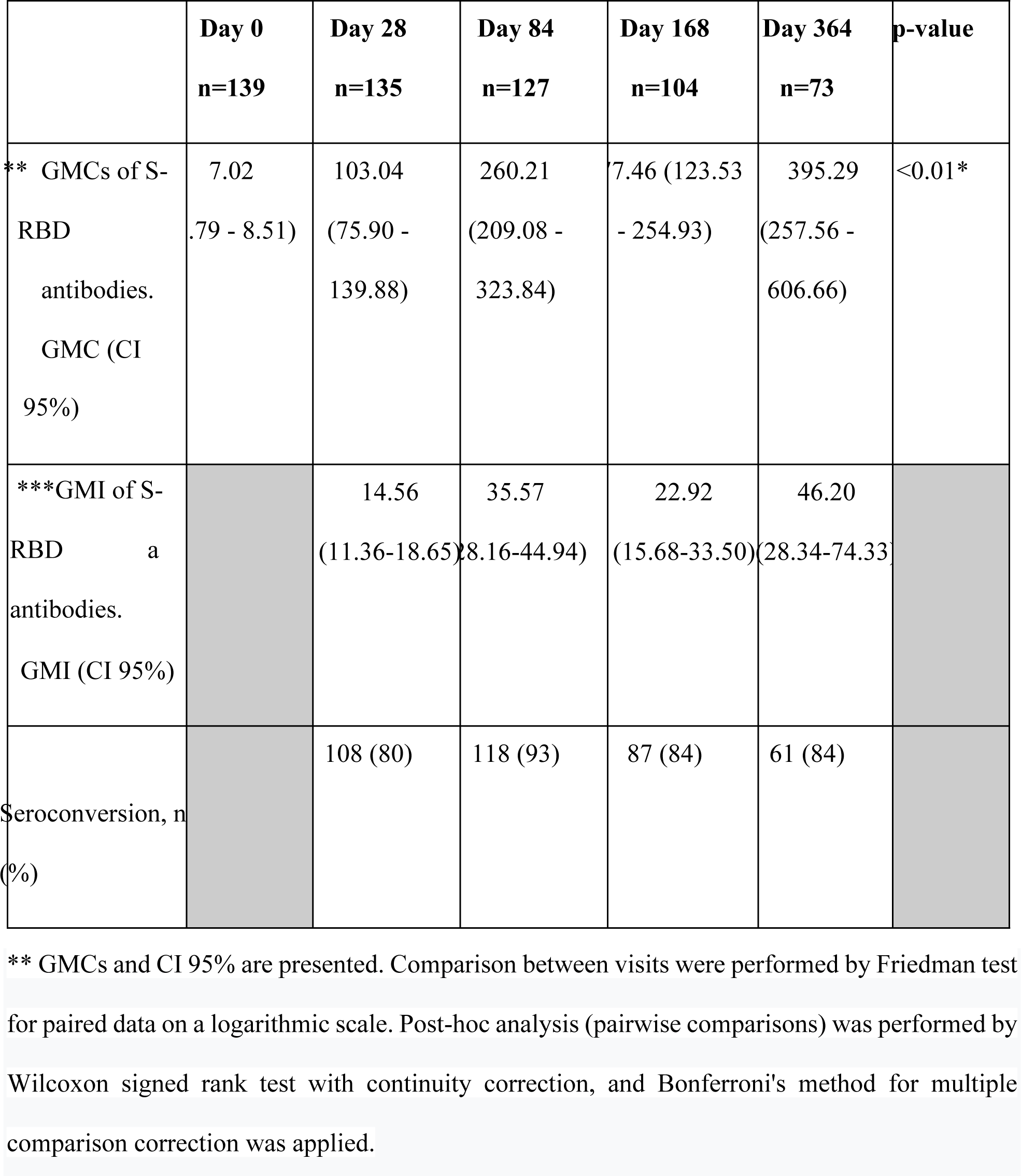
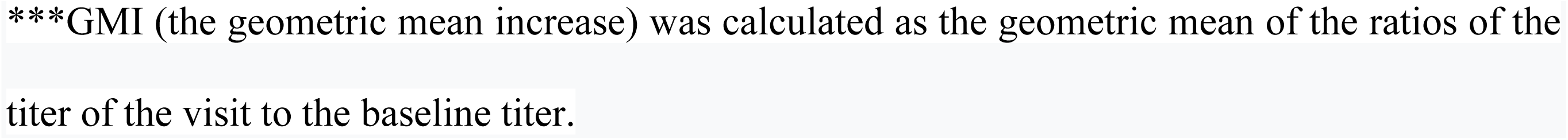
S-RBD antibodies at pre-specified time points after vaccination * *p* < 0.05.

**Table 4.**
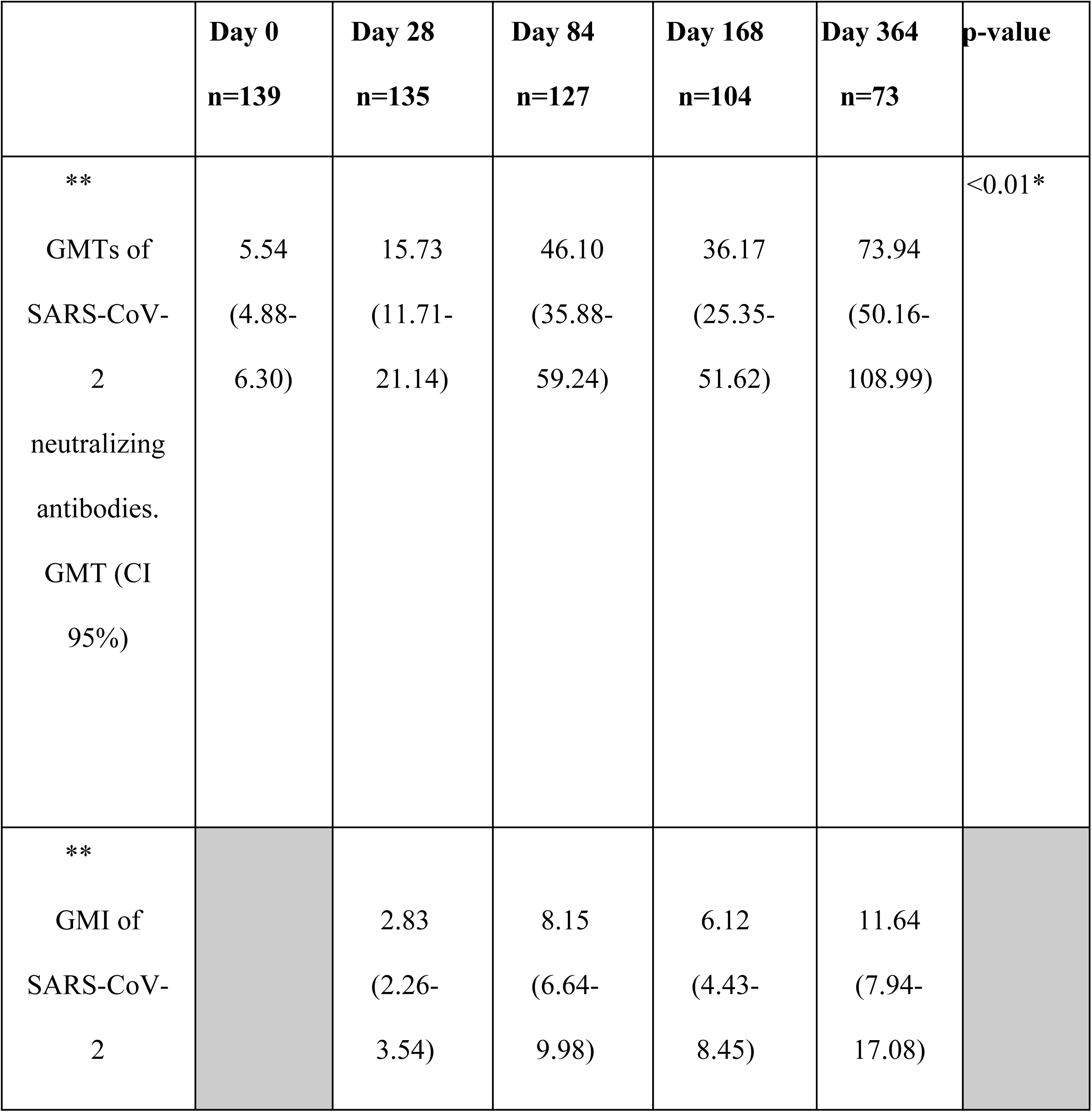
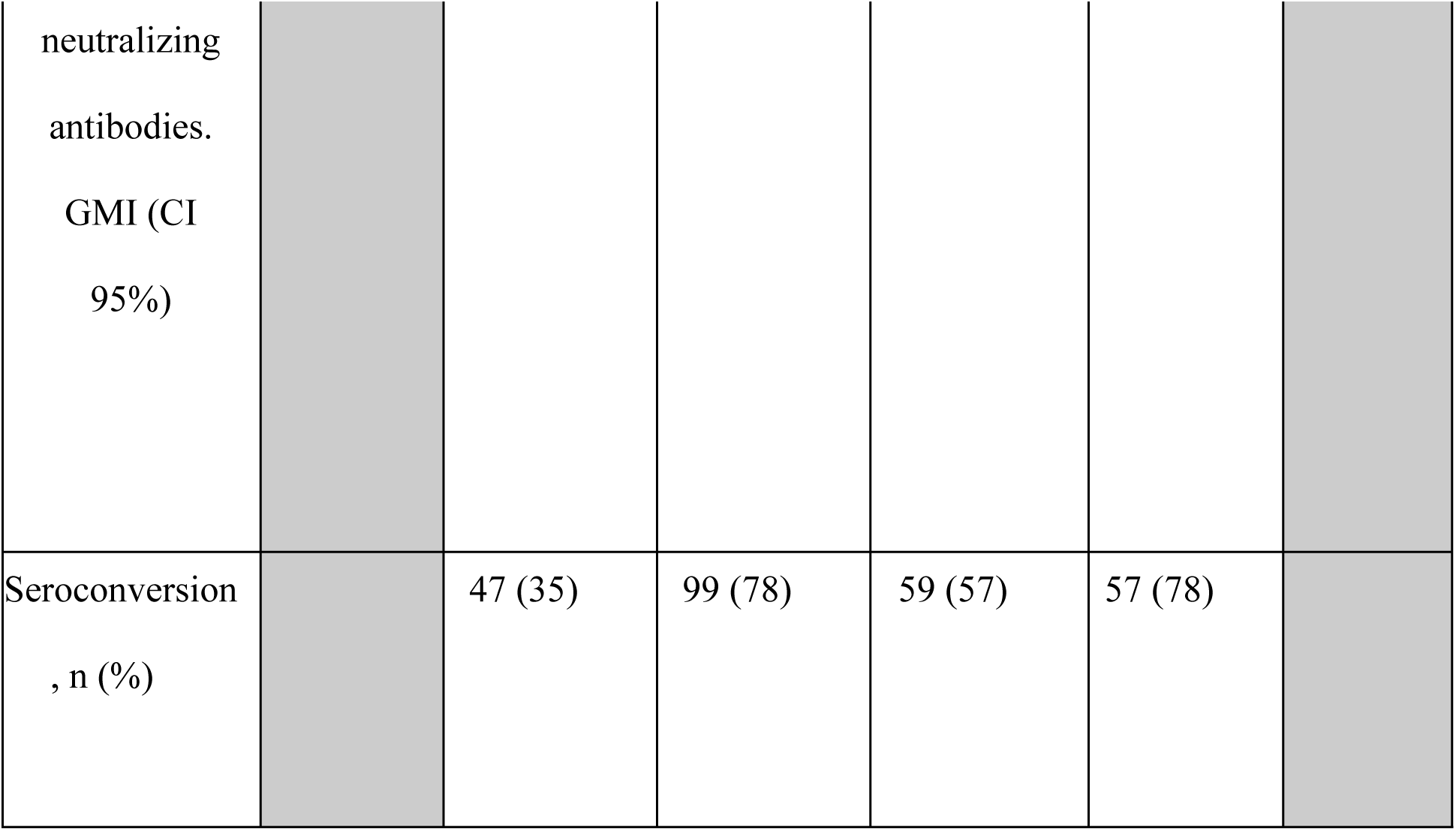
SARS-CoV-2 neutralizing antibodies at pre-specified time points after vaccination.

At baseline, the GMT of S-RBD antibodies was 7.02 (95% Confidence Interval 5.79-8.51), with 38 (27%) participants being categorized as seropositive for S-RBD antibodies. The Ad5-CoV vaccine elicited a strong anti-S-RBD antibody response, which increased after the second dose. There were statistically significant increases in the S-RBD GMT between baseline and all the other visits. Seroconversion of the anti-RBD antibodies occurred in 108/135 (80%) participants on day 28, 118/127 (93%) on day 84, 87/104 (84%) on week 24, and 61/73 (84%) on week 52. Five subjects never reached S-RBD seroconversion.

At baseline, the GMT of nAbs was 5.54 [95% CI 4.88 – 6.30], with 40 (28.1%) participants being categorized as sero-positive for nAbs. nAbs seroconversion occurred in 47/135 (35%) participants at day 28, 99/127 (78%) at day 84; 59/104 (57%) at week 24 and 57/73 (78%) at week 52. There were 21 subjects who did not reach seroconversion of nAbs titers at any time point.

Post-hoc analyses indicated that although there was a significant increase in the GMT of S-RBD antibodies and nAbs in both SARS-Cov-2 sero-positive and sero-negative participants, GMT were significantly higher in sero-positive participants at all time points. Of note, it appears that the GMI rate after the second dose was higher in the SARS-Cov-2 sero-negative group(supplementary figure).

### Exploratory efficacy analysis

There were 45 PCR-confirmed COVID cases among the 133 participants who received two vaccine doses. Of these, 42 (31.6%) occurred within14 days after the second dose: 1 case between 14 and 28 days after the second dose, and 41, between 28 days and 52 weeks. None of the COVID confirmed cases met severe disease criteria. The median time between the application of the second dose and the start date of COVID-19 was 135 days (IQR:81-193). Over half of the COVID-confirmed cases (25/42) occurred in January 2022, when Argentina was in the midst of a new wave of Covid-19 associated with the emergence of the Omicron variant.

## Discussion

Meta-analyses evaluating the immunogenicity and safety of COVID-19 vaccines in PLWH have shown a favorable, though not optimal immune response to COVID-19 vaccines in PLWH[15].

While most studies have shown comparable seroconversion rates to the general population, results on the magnitude of the antibody response have been less consistent with some studies reporting no differences, and other showing lower antibody titers in PLWH. These disparities may be due to the high heterogeneity among studies, including study population, vaccine type, time between vaccine dose and serology testing and definition of seroconversion. Our study adds to the literature by evaluating for the first time an Ad5-nCoV vaccine in PLWH. We found that two doses of the Ad5-nCoV vaccine were well-tolerated and produced an adequate and sustained immune response in PLWH well controlled on ART in Argentina. In addition, the Ad5-nCoV vaccine had a neutral effect on HIV surrogate markers.

In this study, vaccination with Ad5-nCoV had a favourable safety profile with no vaccine-related SAEs, and most solicited AEs being mild in severity. These findings are similar to the previously reported safety profile in the general adult population enrolled in the phase 3 Ad5-nCoV trial[12]. No significant changes in HIV surrogate markers as HIV plasma viral load were found, and CD4, CD8, and CD4/CD8 ratios remained stable across the observation period.

Consistent with prior meta-analysis[15,16], our results indicate a favorable immune response to the Ad5-nCoV vaccine in PLWH. For S-RBD antibodies, there was a GMI of almost 15 fold after the first dose, increasing to 35 fold after the second dose, with sustained GMT S-RBD antibodies for up to a year and seroconversion rates of more than 80% of participants. Development of a neutralising antibody response was lower, with a GMI of 2.8 and 8 fold after the first and second dose. The seroconversion rate after the first dose was only 35%, but increased to more than 75% after the second dose. The immune response to the Ad5-nCoV vaccine in PLWH in this study however, appeared to be lower than that observed after one dose of the Ad5-nCoV in the general adult population, particularly for nAbs. Altogether, these findings suggest that two doses of Ad5-nCoV may be needed in PLWH to achieve adequate protection, given evidence indicating a strong correlation between nAbs and vaccine efficacy against COVID-19[17].

This study has some limitations. First, this was an open-label single arm study, without concurrent recruitment of adults without HIV, and thus post-hoc comparisons with the general population from the phase 3 Ad5-nCoV trial should be interpreted with caution. Second, we included PLWH who were mostly on ART, with well controlled HIV infection and good immunological status. Additionally, our study population predominantly consisted of white men. This may limit the generalizability of our findings to the overall population of PLWH, without previously considering their immunosuppression and ART status, sex or race. Third, although seroconversion rate is a proxy for vaccine efficacy, our study was not powered to evaluate clinical efficacy endpoints and the estimated sample size changed after he rapid advancement of the Argentinean COVID vaccination program. Also a limitation is the number of samples available for immunogenic analyzes through the 52 weeks.

In conclusion, two doses of the Ad5-nCoV vaccine were safe and induced an adequate immune response in virologically suppressed PLWH, maintaining high antibody titers at least during the first year post-vaccination. Future studies should evaluate the clinical efficacy of various immunization strategies with the Ad5-nCoV vaccine in a diverse population of PLWH (including those with more advanced immunosuppression) in preventing breakthrough infections and severe COVID-19 disease by emerging variants of concern.

## Data Availability

All relevant data are within the manuscript and its Supporting Information files

## Contributors

PC, LB, TZ, SH designed the study, and wrote the study protocol. MC,-GM, AN oversaw statistical analyses. PC, MIF, GL, IC, MJR, and MY accessed and verified the underlying data in the manuscript. LG and the quality team led the implementation and study quality assurance. All authors had full access to all the data in the study and had final responsibility for the decision to submit for publication. All authors approved the final manuscript.

## Declaration of interests

PC received grant support to his institution from CanSino Biologics Inc. To carry out this study as an investigator initiative, Fundacion Huesped was responsible for site selection, study implementation, quality control and assurance, and general overview of the study. Other authors declare no competing interests.

## Data sharing

Fundacion Huesped and the Canadian Center for Vaccinology shared anonymised individual patient data with Cansino Biologistic Inc.

Role of the funding source

CanSino Biologics provided the vaccine and placebo, as well as the funds for the study.

## Acknowledgements

We want to acknowledge the invaluable and generous contributions of all study participants and their families, and of all the staff across the participant sites who supported recruitment and retention into the study. We acknowledge M. Eugenia Socias for her contribution to the writing of this manuscript, Javier Mariani and team for the statistical analysis, and Meixu Yan for the contribution, support and work on the manuscript.

## Notes

### Competing Interest Statement

I have read the journal's policy and the authors of this manuscript have the following competing interest: Fundacion Huesped reports financial support was provided by CanSino Biologics Inc. Pedro Cahn reports financial support was provided by Fundacion Huesped. Pedro Cahn reports a relationship with ViiV Healthcare that includes: consulting or advisory. Pedro Cahn reports a relationship with Gilead Sciences Inc that includes: consulting or advisory. Patricia Patterson reports a relationship with Host Foundation that includes: employment. PATRICIA PATTERSON has patent MD pending to 84274. Dr. Luis Barreto reports financial support and administrative support were provided by CanSino Biologics Inc. Dr. Luis Barreto reports a relationship with CanSino Biologics Inc that includes: consulting or advisory and travel reimbursement. Member of the Scientific Advisory Board of CanSino Biologics Inc. No conflict of interest If there are other authors, they declare that they have no known competing financial interests or personal relationships that could have appeared to influence the work reported in this paper.

### Clinical Trial

ClinicalTrials.gov:NCT05005156

### Funding Statement

Yes

### Author Declarations

iniciativa y Reflexión Bioética -IRB- approval number: 3978. informed consent was taken in written form

